# Exploring relationships between drought and epidemic cholera in Africa using generalised linear models

**DOI:** 10.1101/2021.07.16.21260629

**Authors:** Gina E C Charnley, Ilan Kelman, Nathan Green, Wes Hinsley, Katy A M Gaythorpe, Kris A Murray

**Author notes:** Gina E C Charnley (corresponding author), MSc, School of Public Health, Imperial College London, Norfolk Place, London, W2 1PG.

## Abstract

**Background:** Temperature and precipitation are known to affect *Vibrio cholerae* outbreaks. Despite this, the impact of drought on outbreaks has been largely understudied. Africa is both drought and cholera prone and more research is needed in Africa to understand cholera dynamics in relation to drought.

**Methods:** Here, we analyse a range of environmental and socioeconomic covariates and fit generalised linear models to publicly available national data, to test for associations with several indices of drought and make cholera outbreak projections to 2070 under three scenarios of global change, reflecting varying trajectories of CO_2_ emissions, socio-economic development, and population growth.

**Results:** The best-fit model implies that drought is a significant risk factor for African cholera outbreaks, alongside positive effects of population, temperature and poverty and a negative effect of freshwater withdrawal. The projections show that following stringent emissions pathways and expanding sustainable development may reduce cholera outbreak occurrence in Africa, although these changes were spatially heterogeneous.

**Conclusions:** Despite an effect of drought in explaining recent cholera outbreaks, future projections highlighted the potential for sustainable development gains to offset drought-related impacts on cholera risk. Future work should build on this research investigating the impacts of drought on cholera on a finer spatial scale and potential non-linear relationships, especially in high-burden countries which saw little cholera change in the scenario analysis.

## Background

*Vibrio cholerae* is a water-borne bacterial pathogen, causing profuse watery diarrhoea and rapid dehydration in symptomatic cases. This can lead to death within two hours of symptom onset and case fatality ranging from 3-40% [1,2]. The seventh and ongoing cholera pandemic began in 1961, spreading to Africa by the 1970s, where it now shows signs of endemicity in several countries [3,4]. Despite over 94% of World Health Organization (WHO) reported cholera cases occurring in Africa and some of the highest mortality rates [5], previous research has heavily focused on South America, the Indian subcontinent and more recently Haiti.

Cholera outbreak frequency is closely related to environmental and climatic changes [6,7,8]. For instance, temperature and precipitation are considered important in cholera outbreak occurrence, with temperature driving epidemics and precipitation acting as a dispersal mechanism [9]. These relationships have implications for cholera outbreaks after natural hazards, such as droughts. Several links between drought and cholera outbreaks have been described [2,10,11] and it is hypothesised that increasing concentrations of infectious bacteria in more limited drinking water sources and increased risky drinking water behaviours are likely mechanisms for transmission [2,12]. Despite this, drought and cholera in Africa are understudied in isolation and links have more commonly been made between flooding, despite droughts potentially posing a considerably greater risk than floods [11].

Cholera is considered a disease of inequity and several socio-economic risk factors have been implicated with cholera outbreaks, which may be further exacerbated by droughts. Some studies suggest that human-induced factors are more important for cholera dynamics than climate or environmental ones [13], including poverty [14], sanitation [15], drainage [16], water quality [17] and poor healthcare [9]. This supports the notion that outbreaks result from the breakdown of societal systems responses to a hazard, leading to a human-environment link and subsequent pathogen shedding [18]. Water, sanitation and hygiene (WASH) factors are considered particularly significant, as the importance of the water body reservoirs depends on the sanitary conditions of the community [19]. Eight hundred and forty-four million people worldwide lack access to basic drinking water and a further 2.4 billion are without basic sanitation [20], putting many people at risk of water-borne disease outbreaks including cholera.

Here, we aim to address the research gap of drought-related health outcomes by investigating its implications on cholera. The work fills an important research gap as few studies have investigated the link between drought and cholera outbreaks in Africa, or projected outbreak changes into the future, and investigating mechanisms through which global change might yield health impacts. Research in this area is particularly important due to a significant number of people at risk of both cholera and drought and the negative implications that climate change may have for these communities.

## Methods

In this study, we aimed to understand the implications of drought for cholera outbreak occurrence at a continental scale across Africa, after accounting for important socio-economic factors. We aim to use these results to further understand the hypothesis that droughts lead to cholera outbreaks through elevated pathogen concentrations in limited water and an increase in risky drinking water behaviours, Fig. 1 shows a schematic to help visualise this hypothesis and potential pathways. In addition, we aimed to evaluate how future changes in drought area and risk due to climate change [21,22], alongside other development factors, may impact future cholera outbreak occurrence. We thus developed several projection scenarios incorporating different greenhouse gas emissions and socio-economic development trajectories.

**Fig. 1.**
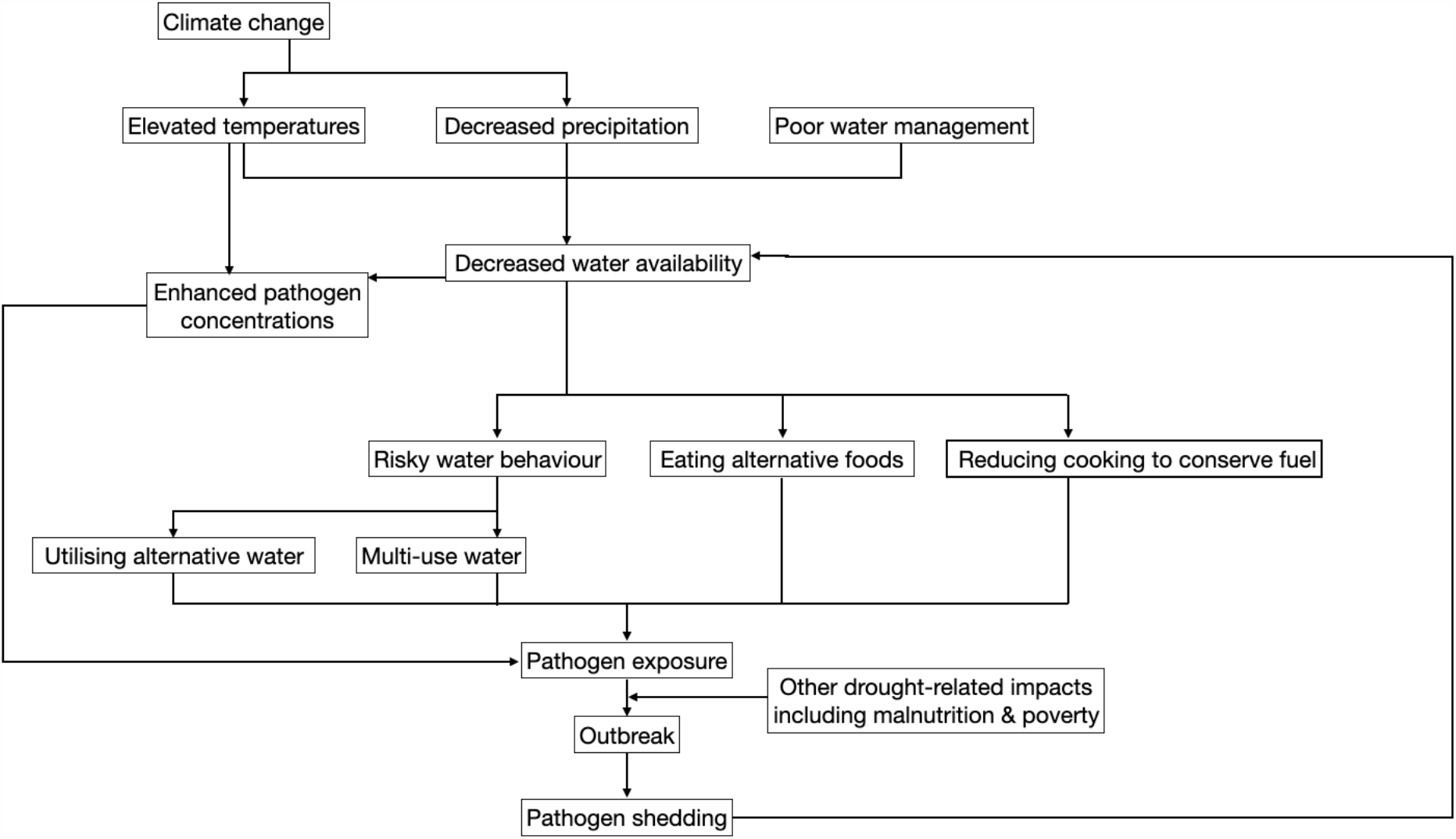
Pathways from water shortages to cholera outbreaks: Suggested mechanism through which drought can lead to cholera outbreaks in Africa [2,12]

### Datasets and Study Period

We compiled data on cholera outbreaks and a range of social and environmental covariates over the period 1970 to 2019. Annual cholera cases were retrieved from the WHO’s Global Health Observatory [23], which provides reported annual cholera case for each country, which were confirmed either clinically, epidemiologically, or by laboratory investigation. For analysis, these numbers were transformed into a binary outcome to reflect outbreak occurrence (i.e., set at 0 for no outbreak and 1 for an outbreak (>1 case/death)), which was then used as the outcome variable in the models. We opted not to analyse raw case data to minimise the effect of unmeasured observations and reporting biases among countries. For years with no outbreak data, the outcome was set to 0, assuming if cholera cases/deaths occurred within a country then they would have been identified and reported (a sensitivity analysis for this assumption is presented in Supplementary Information 1).

In total, 19 environmental and socio-economic covariates were selected for investigation based on prior hypotheses and previous results linking cholera outbreaks to risk factors (summarised in Supplementary Table 1). Environmental data were extracted from a variety of sources and included climate (temperature and precipitation) [24], meteorological drought (Palmer Drought Severity Index, PDSI) [25], agricultural drought (soil moisture and potential evapotranspiration (PET) [26,27] and hydrological drought (runoff and freshwater withdrawal annually and per capita) [28,29]. Where monthly or sub-national data were available, we calculated national yearly means. Climate data were missing for Côte d’Ivoire and drought data were missing for Rwanda, The Gambia, Guinea-Bissau, Djibouti, Burundi, Benin, Cabo Verde, São Tomé and Principe, Comoros, Mauritius and Seychelles.

Environmental data for these countries were derived by taking the mean of their neighbouring countries, whereas islands were excluded.

Socio-economic data including annual indicators of poverty and development, WASH, malnourishment, and population (on a logarithmic scale), were taken from the WorldBank [30] and the United Nations Development Programme [31] datasets. Where a country’s socio-economic data were missing for some years, a national average was taken from the available data points and used for all years. If national data were missing for the full instrumental period, these countries were removed from the analysis.

After examining data completeness across the full dataset, we designated the instrumental period for analysis to be 2000 to 2016 to limit omitting missing data and interpolation. Summary figures of the climate and cholera data over the instrumental period are shown in Supplementary Figure 1. Summary figures of the drought indices and their definitions are shown in Supplementary Figure 2 and Supplementary Information 2.

### Model Structure and Fitting

Generalised linear models (GLM) were fitted to the dataset describing the cholera outbreak occurrence for the instrumental period (2000-2016), for all countries in mainland Africa and Madagascar, using maximum likelihood estimation. Due to the binary outcome variable for cholera outbreak occurrence, a binomial likelihood with a log-log link function was used in all models. Rows with missing values were removed from the data frame.

From this initial dataset, a reduced pool of potential covariates was selected for model fitting using a covariate selection process developed by Garske *et al*. [32] and Gaythorpe *et al*. [33]. In summary, univariate models for each potential variable were fitted to the binary outcome variable and any variables not significantly associated with the outcome at a 10% confidence limit (p<0.1) were excluded. Of the remaining covariates, absolute pairwise correlations were calculated, and highly correlated variables (r>0.75) were then clustered into groups. The covariates from each cluster most strongly correlated with the outcome variable was then selected for inclusion in the multivariate models, fit using the function glm. Model fit was evaluated using Bayesian Information Criterion (BIC) and a single best-fit model was found using the stepAIC function. In addition, area under the receiver operator characteristic curve (AUC) was used to quantify model performance. All statistical analyses were carried out in R Studio version 3.6.2 (packages: tidyr, MASS, ggplot2, dplyr, magrittr, corrplot, caret, nlme, MuMIn, car, boot).

### Testing for Temporal and Spatial Effects

The inclusion of multiple years of data across multiple countries raises the possibility of spatial and temporal confounding (e.g., autocorrelation). To investigate the potential influence on the covariate selection and subsequent model, separate analyses were run including year and ISO3 country code as predictor variables following the same step-wise covariate selection process and multivariate model approach as described above.

Autocorrelation diagnostics were run on selected spatial and temporal covariates by testing the significance of the linear relationship with and without consideration of AR1 (autoregressive model of order 1) autocorrelation and assessing evidence of autocorrelation in the residuals. Leave-one-out (LOO) cross validation using Akaike Information Criterion (AIC) was used to assess model performance of both the original (without year/ISO) and the updated (with year/ISO) multivariate models selected through the covariate selection process.

### Projection Scenarios

Three scenarios (S1, S2 and S3) were developed for 2020-2070 (at decadal increments) as summarised in Table 1. Each scenario represents an alternative possible future trajectory of the variables retained in the best fit model, parameterised to varying degrees of climate mitigation and socio-economic development. Here, S1 represents a “best-case” scenario, loosely aligning to highly ambitious climate change mitigation and strong progress towards the Sustainable Development Goals (SDG), S2 represents an intermediate scenario, and S3 a “worst-case” scenario with slower progress towards emissions reductions and the SDGs.

**Table 1.**
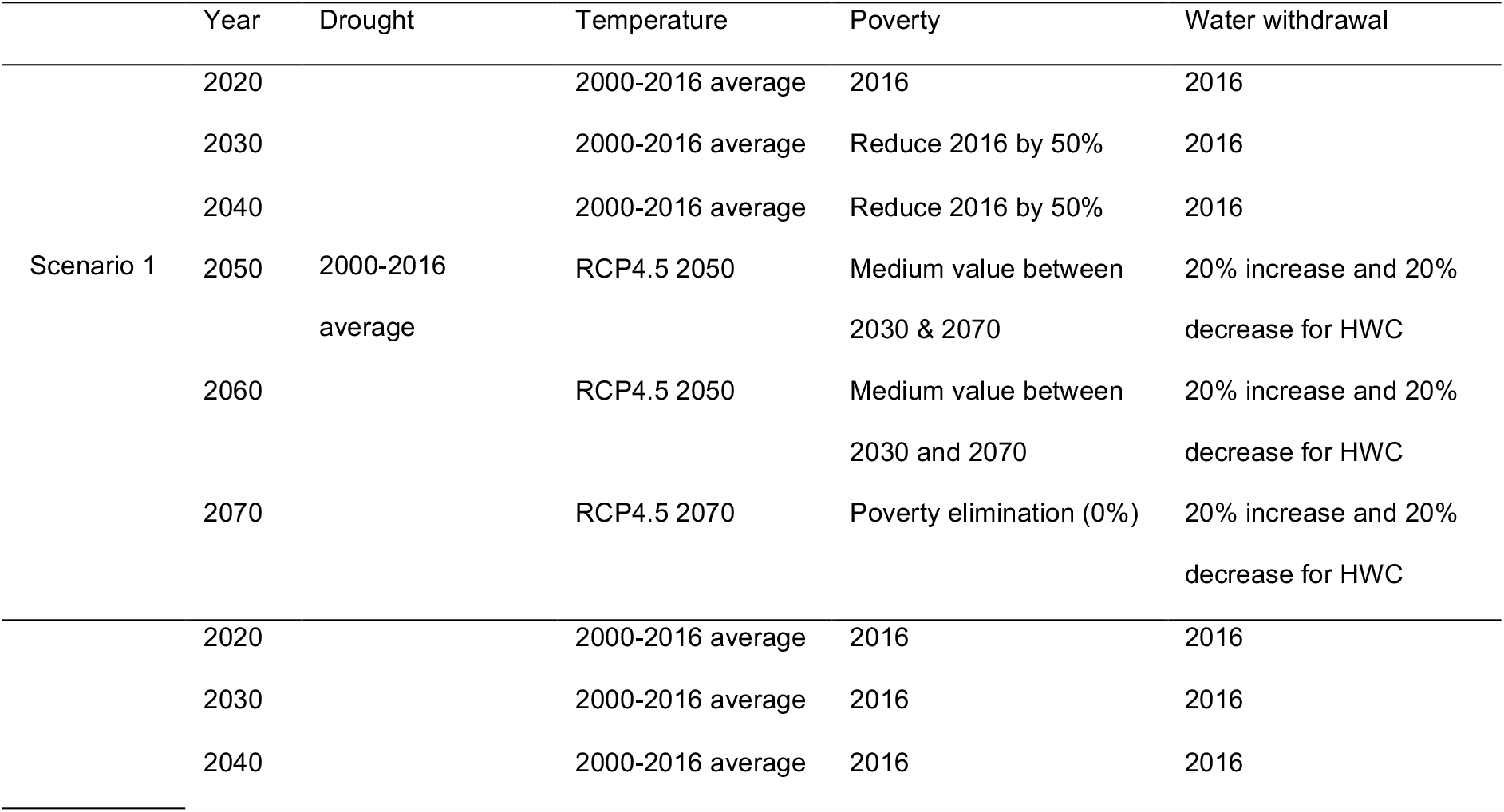
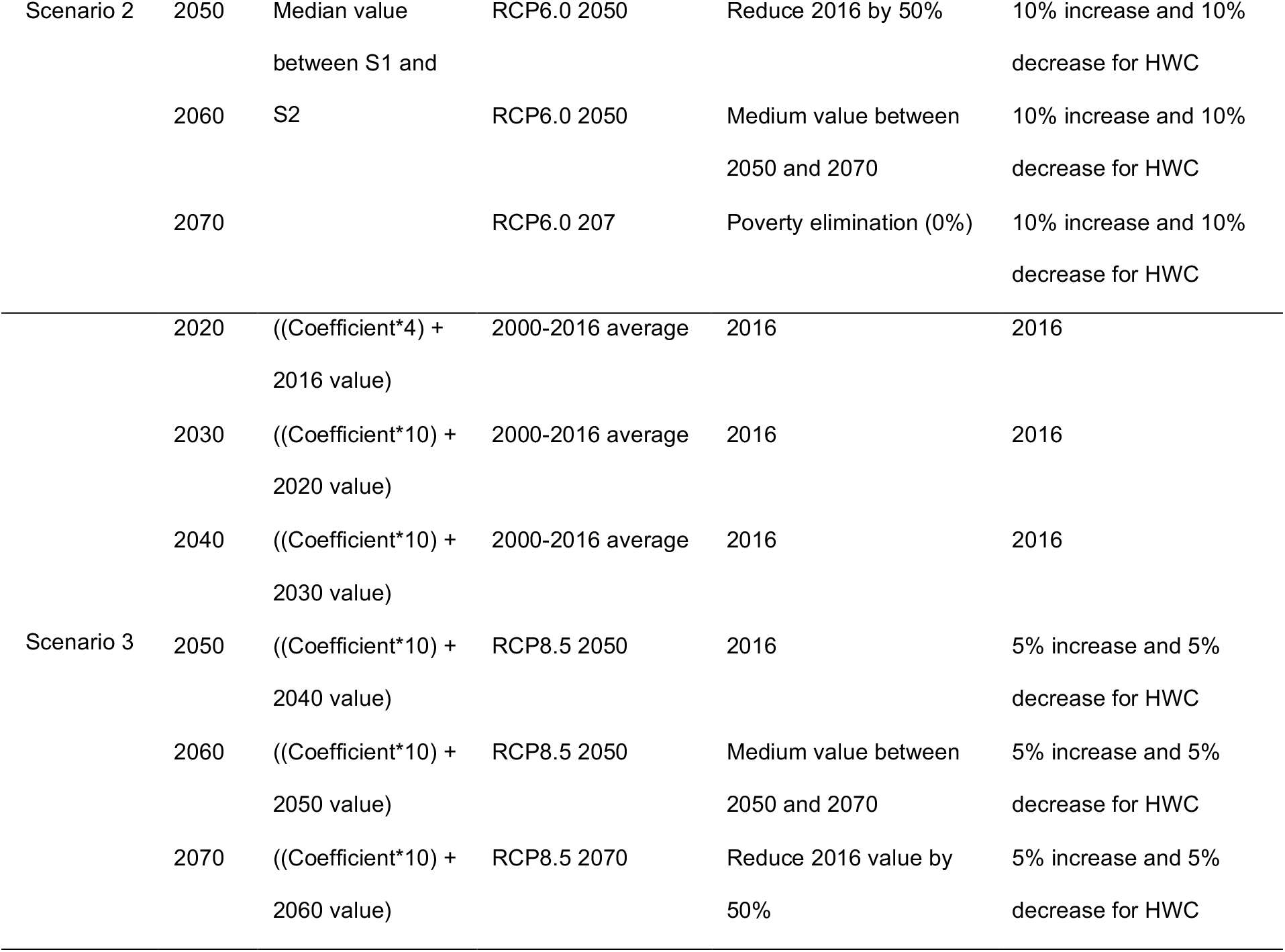
Cholera projection scenarios for 2020-2070 at decadal intervals: Scenario 1 (S1), a “best-case” scenario; Scenario 2 (S2), an intermediate scenario and Scenario 3 (S3), a “worst-case” scenario. The scenarios were projected over fifty years from 2020-2070. HWC = high withdraw countries including MDG, LBY, SDN, MRT and MAR

Detailed descriptions and justifications of the projected changes for each variable are provided in full in Supplementary Information 3. Briefly, projected temperature data (for 2050 and 2070) were taken from WorldClim [34], as this was also used for historical data. The data is Coupled Model Intercomparison Project 6 (CMIP6) downscaled future climate projections, processed for nine global climate models using three Representative Concentration Pathways (RCP). We used RCP4.5, 6.0 and 8.5 for scenarios S1, S2 and S3, respectively. This was projected for 2050 and 2070 and we used the instrumental period average (2000-2016) for 2020-2040 values. The average was used to account for interannual climate variability. Supplementary Figure 3 summaries the data for each pathway and year.

Projecting PDSI at a continental or national scale is contentious showing a range of projection outcomes, due to high spatial heterogeneity and between model uncertainty/disagreement [21,35], as well as computational discrepancies depending on the PET algorithm used [*36*]. Several PDSI modelling studies [36,37] and paleoclimatic studies [38,39] found that drought severity and durations remained constant despite periods of extreme dryness, over a range of time scales. We also observed this in our dataset for both the full data range and the instrumental period and our data accurately captured past drought as its changes tracked with soil moisture, a good index of drought (see Supplementary Figure 4 and 5) [41]. Given these disagreements and following other drought projection studies [40], we opted to estimate future drought conditions for each scenario as follows: For S1, we included no change relative to a current “baseline” by fixing drought values to the instrumental period average (2000-2016), the average was used to account for interannual climate variability. For S3 (representing “business-as-usual”) we extrapolated the full historical data trends for each country (1850-2016) using univariate linear regression models (drought∼year). The results of these models are available in Supplementary Table 2 and the coefficients then acted as a yearly multiplier (up until the extreme values of +4 for extreme wetness and -4 for extreme dryness). For S2, we took an intermediate value between S1 and S3. To account for uncertainty in the drought projections and to further examine how drought in isolation may alter future cholera outbreaks, a second sensitivity analysis was run, maintaining the other covariates at the 2016 levels and altering drought in six analyses +/-0.5, +/-1 and +/-2 (or until the extreme values, +4 or -4). Full details and results of this sensitivity analysis are shown in Supplementary Table 3 and Supplementary Figure 6.

Poverty changes were based on SDG 1 [42], despite the limitations of the SDGs (e.g., ambiguous terms), they are a globally recognised standard for sustainable development. As such, S1 meets the goal of a 50% reduction in extreme (<$1.25/day) poverty by 2030 and poverty eliminated by 2070. In S2, the 50% reduction goal is met by 2050 and by 2070 for S3. The poverty setting used in the SDGs is slightly lower ($1.25) than the WorldBank data used in this analysis ($1.90), and it is difficult to distinguish the level of poverty within the data; therefore, the projected scenarios mainly aligned with the second part of the goal, to halve the population in poverty by 2030.

Projected changes in freshwater withdrawal are largely dependent on future human behaviour and adaptation to changing water security, which are highly uncertain. Therefore, freshwater withdrawal projections were based on SDG6.4 and either increased or decreased based on each country’s historical freshwater withdrawal relative to available water resources, taken from the same source used in the model [28]. This indicator of freshwater security for each country is plotted in Supplementary Figure 7. Expanded freshwater withdrawal would likely increase peoples’ access but this must be done sustainably and in line with resources. Increased withdrawal may also be a sign of development as more people have access to wells, boreholes and piped water. As such, for S1 we increased sustainable freshwater availability by the middle of the projection period (2050) by 20% for countries with sufficient resources. For S2, we increased freshwater availability by 10% and for S3 by 5%.

For population projections, the United Nation’s World Population Prospectus [43] median variant was used for all three scenarios. Although population growth is expected to be more restricted under high attainment of the SDGs, we opted to use a single medium population size to isolate the effects of the other environmental and socio-economic covariates.

## Results

### Model Fitting and Covariate Selection

The univariate model results (p-values, coefficients, BIC and AUC of the 19 tested covariates against cholera outbreak occurrence) are shown in Table 2. Six of these were not significantly associated with the data at the threshold of p<0.1. Of the remaining 13, one cluster was formed containing two highly correlated variables (soil moisture and drought), while all other covariates were considered uncorrelated at the given threshold and therefore could be included in the full model.

**Table 2.**
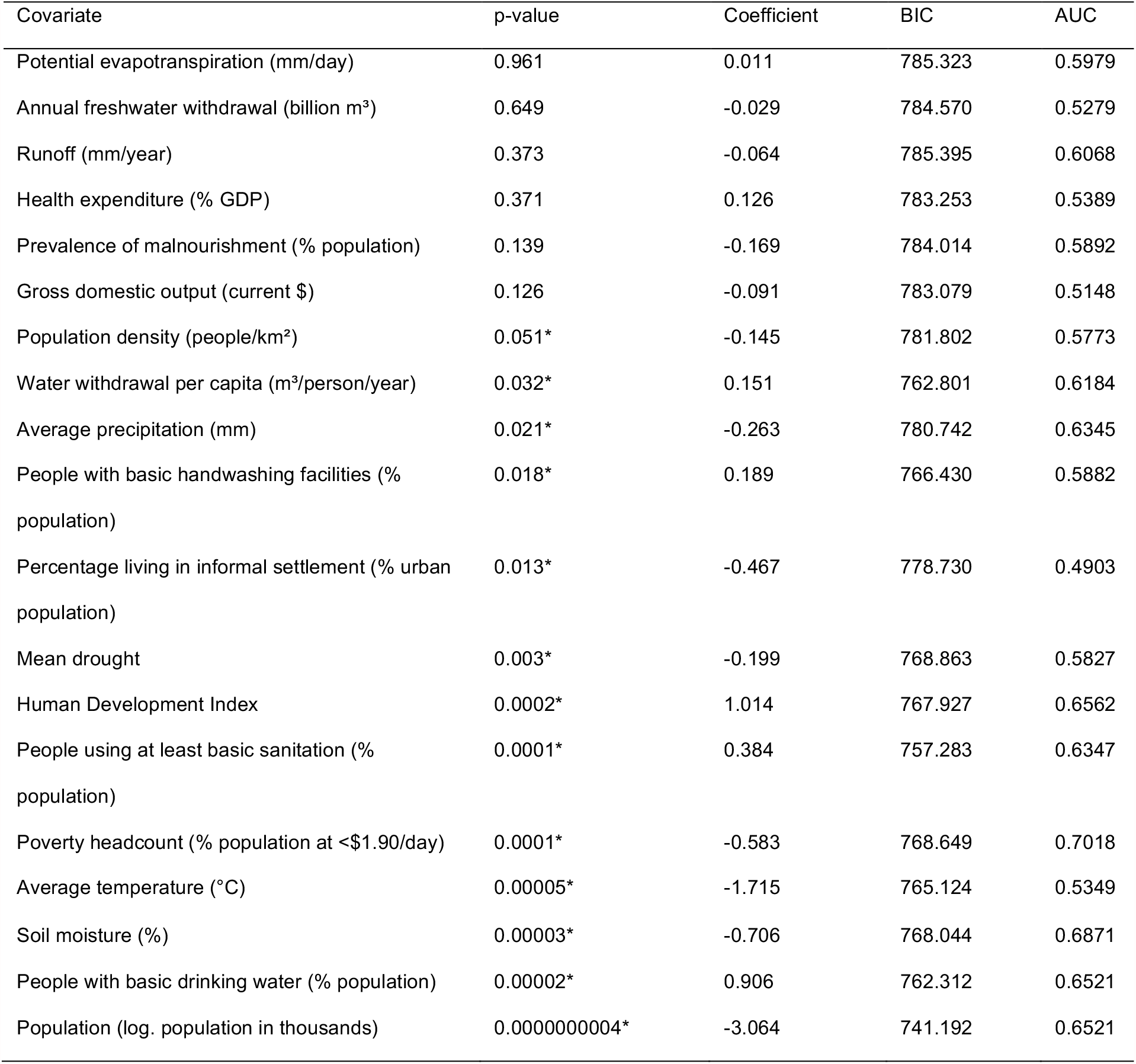
Univariate model outputs and goodness-of-fit measures for the tested covariates against cholera outbreak occurrence, including p-values, coefficients, BIC and AUC. ^*^ p<0.1

### Output from the Best-Fit Model

After model fitting, five covariates were retained in the best-fit model. These include population, mean meteorological drought (in PDSI), average temperature, poverty headcount and per capita freshwater withdrawal. Goodness of fit measures and outputs for the best-fit model are shown below in Table 3. Higher population numbers and more people living in poverty were associated with increased cholera outbreaks. For the environmental covariates, per capita freshwater withdrawal was negatively associated with cholera, while higher temperatures and drier conditions (more negative PDSI) were both associated with increases in cholera outbreaks. These relationships are shown in the marginal effect plots in Fig. 2.

**Table 3.** Output and goodness of fit measures for the best-fit model

|  |  |  | Coefficient | Exp(Coefficient) | p-value |
| --- | --- | --- | --- | --- | --- |
| Mean national drought (PDSI) |  |  | -0.0927813 | 0.9113928127 | 0.051172 |
| Population, total (log) |  |  | 1.3125412 | 3.7156036497 | 2.85x10<sup>-13</sup> |
| Average temperature (°C) |  |  | 0.0927423 | 1.0971789754 | 0.000113 |
| Poverty headcount (at <\$1.90/day) | | | 0.0327487 | 1.0332908900 | 4.23x10<sup>-16</sup> |
| Per capita freshwater withdrawal (m<sup>3</sup>/person/year) |  |  | -0.0024225 | 0.9975804550 | 5.43x10<sup>-7</sup> |
| Residuals | Min | 1Q | Median | 3Q | Max |
|  | -2.0286 | -0.7974 | 0.4069 | 0.8601 | 2.2564 |
| R<sup>2</sup>: 0.276254 |  |  |  |  |  |
| AUC: 0.7784 |  |  |  |  |  |

**Fig. 2.**
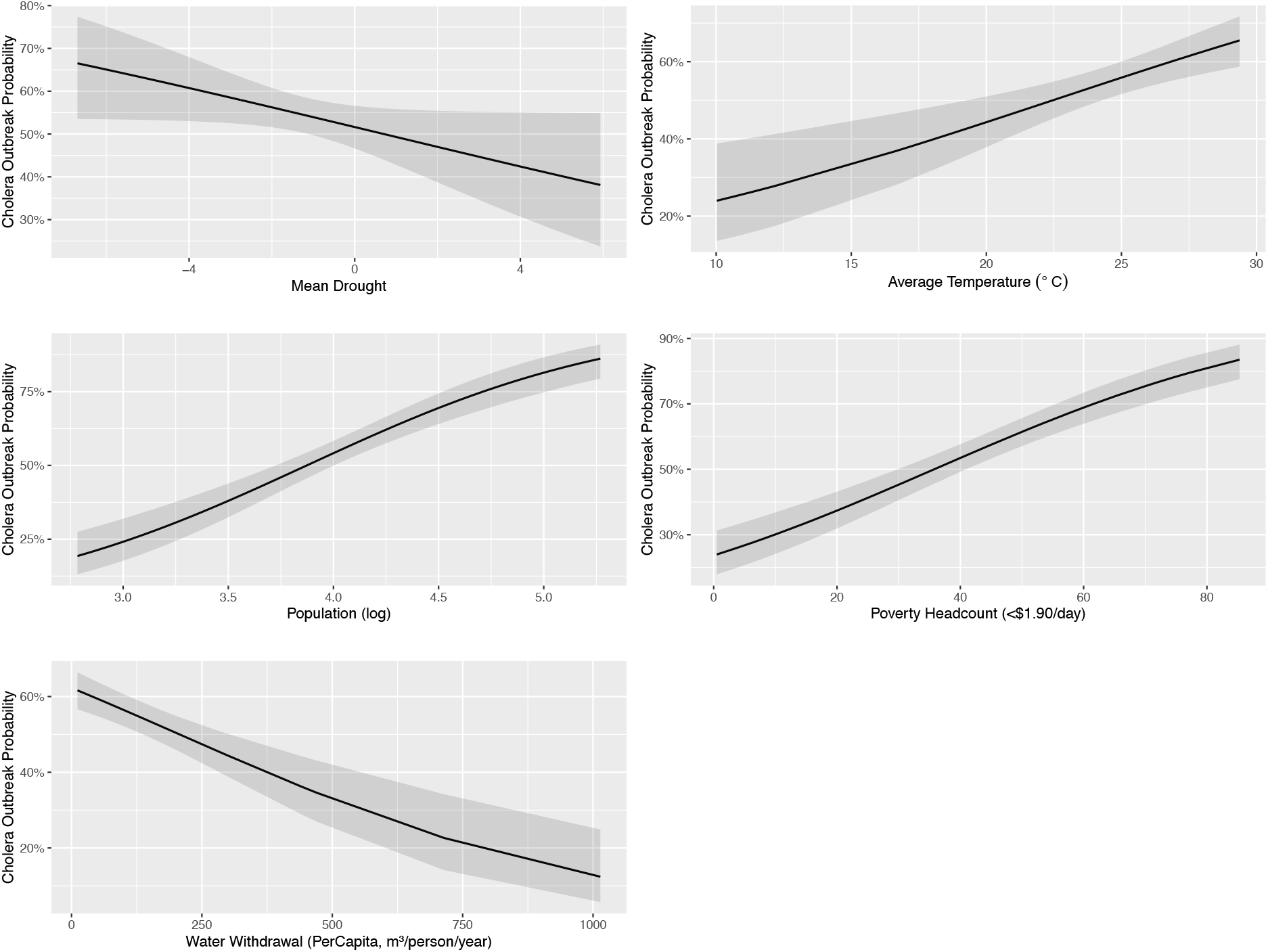
Marginal effect plots for the five selected covariates for the best-fit model, showing cholera outbreak occurrence probability

### Temporal and Spatial Effects

Re-running the covariate selection process with year, ISO3 country code and the 19 original predictor variables, selected year but not ISO3 at the significance threshold (p=<0.1). It also selected the same covariates as the original model and additionally basic handwashing and Human Development Index. The linear relationship between year and cholera was visualised using loess curves for each country (shown in Supplementary Figure 8) and when accounted for AR1 autocorrelation year was found to no longer be significant (p=<0.05).

Out-of-sample validation using AIC and LOO found no appreciable difference between the two selected best-fit models. Therefore, the model selected without the inclusion of year and country code in the selection process was thus as the best-fit model (diagnostic results are shown in Supplementary Information 4).

Cholera outbreak occurrence appears conditionally independent of year given the other covariates in the model, as time does not cause cholera but instead the changes in covariates over time, making them good predictors of cholera outbreak occurrence. It is also thought that some temporal increases in cholera are due to global improvements for detection of all-pathogen outbreaks from the mid 1990s onwards, especially in low- and middle-income countries, improving countries’ capacity for detection, response and therefore reporting [44,45].

### Cholera Projections to 2070

Cholera projections from the best-fit model according to the parameter values for each of the three scenarios are shown in Fig. 3. The cholera outbreak projections show several changes through to 2070 and spatial heterogeneity among countries over the continent. Most countries show a general decrease in cholera outbreaks in S1 and S2, with few exceptions e.g., Tunisia. Although countries with the highest cholera levels saw little change, remaining at a high outbreak occurrence level throughout, including the Democratic Republic of Congo (DRC) and Nigeria.

**Fig. 3.**
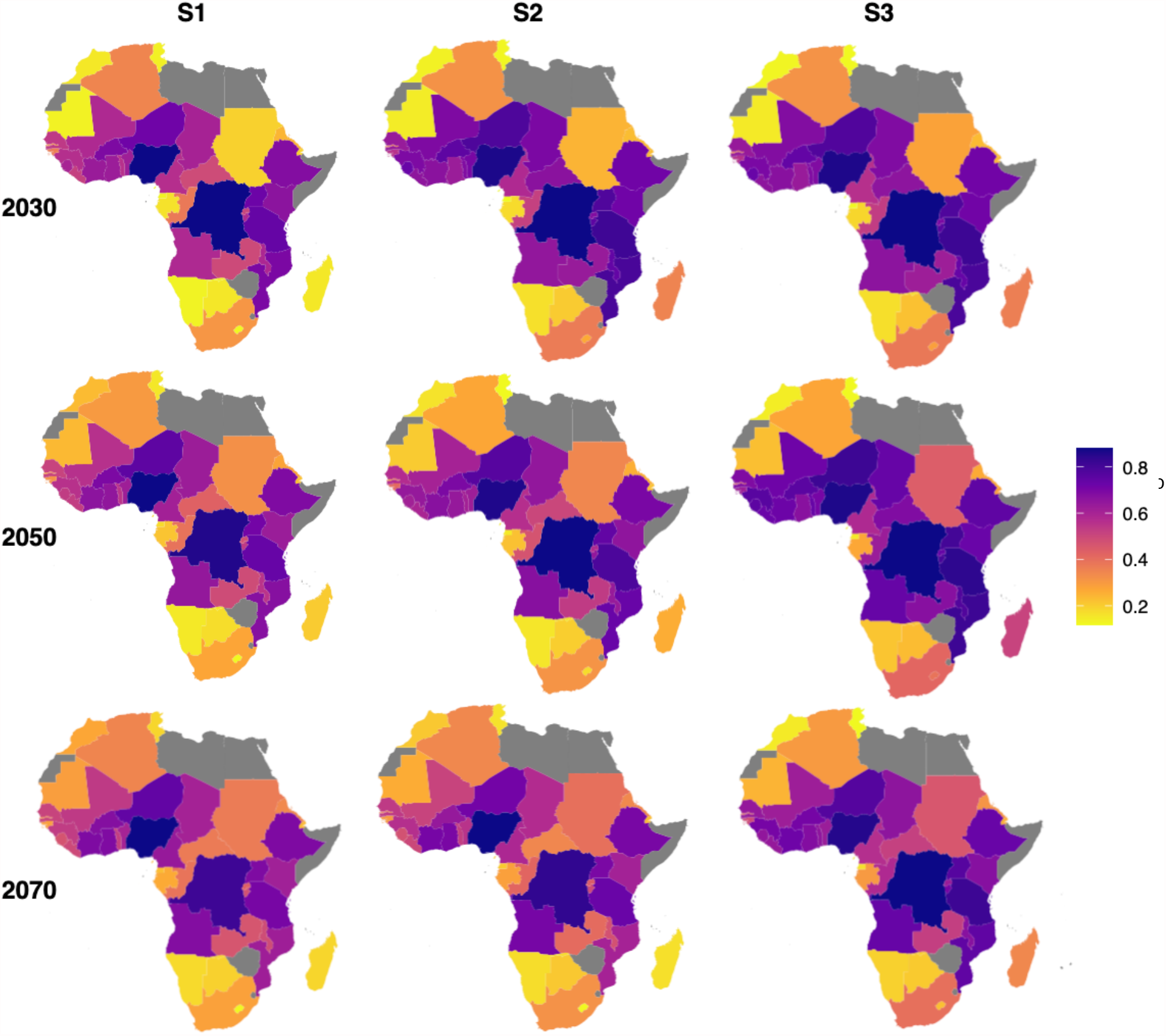
Projected cholera outbreak occurrence (0-1) for the three scenarios in 2030, 2050 and 2070. Grey represents countries where covariate data was missing (Botswana, Zimbabwe, Somalia, Egypt, Eswatini, Western Sahara, Algeria, Libya and Eritrea) and therefore could not be included in the model

Fig. 4 shows the decadal continental average for the projected cholera outbreak occurrence, to help understand the general trend across the continent. Overall, S3 shows a slight increase throughout the projected period, whereas S1 and S2 exhibit declines.

**Fig. 4.**
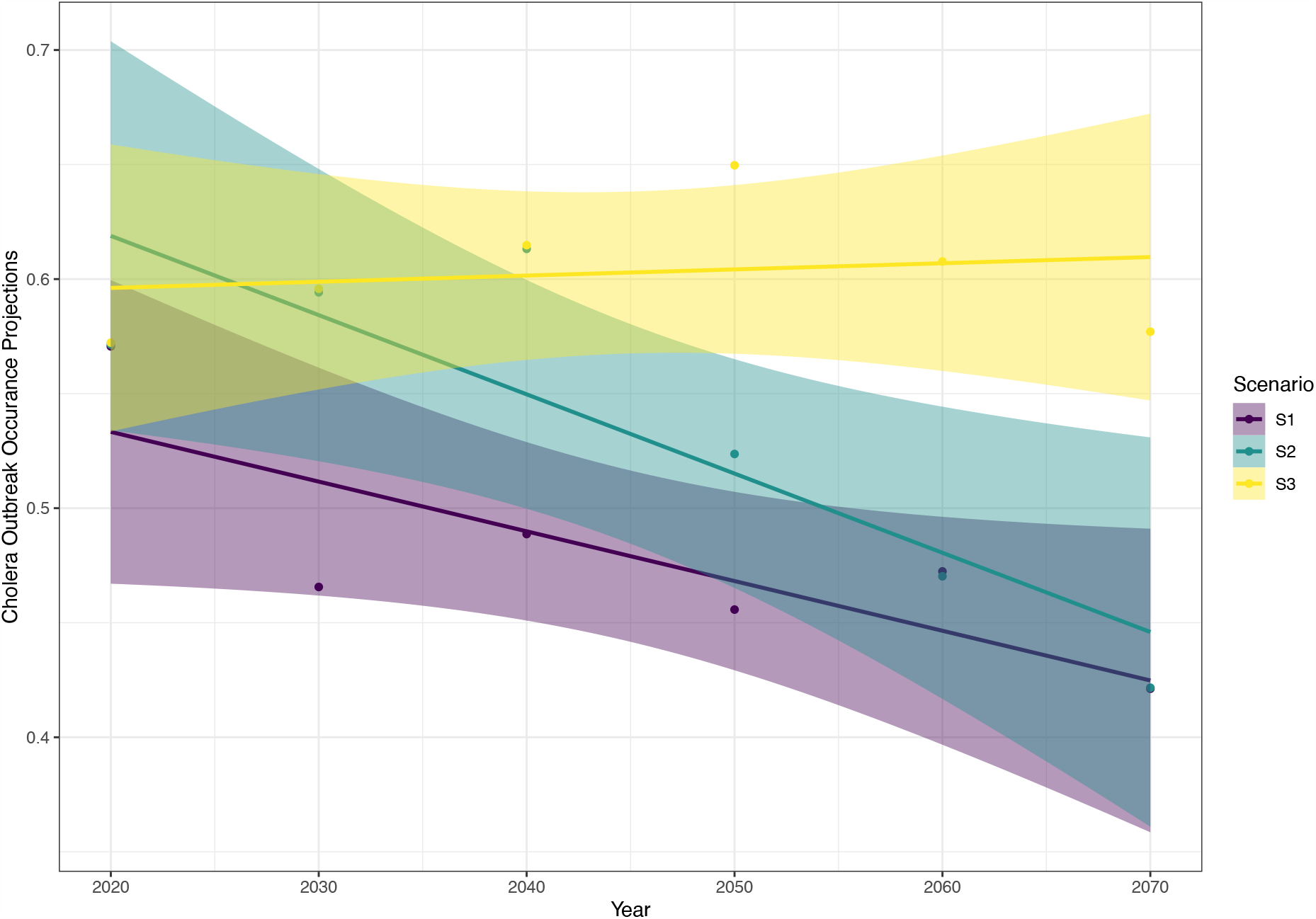
Mean continental cholera outbreak occurrence for the projected period (2020-2070) using the three scenario datasets

However, overlapping confidence intervals between S1 and S2 mean it is difficult to distinguish meaningful differences, although by 2070 S3 projects significantly more outbreaks than S1 and S2.

The drought sensitivity analysis showed modest changes through the six different analyses, with more negative values of PDSI seeing higher cholera outbreak occurrence (Supplementary Table 3 and Supplementary Figure 6). Despite this, these changes were not excessive with a 0.06 average increase in continental cholera outbreak occurrence from the 2000-2016 averages to sensitivity analysis 6 (2016 value – 2). This suggests that while future drought is likely to continue to affect cholera in Africa, improved socio-economic conditions may counteract this effect, by reducing pathogen exposure.

## Discussion

Cholera has well established environmental [6,7,8] and socio-economic links [9,14,15], such as poverty, poor WASH conditions, the Intertropical Convergence Zone and El Niño Southern Oscillation. Here, environmental variables were important covariates in the model. Meteorological drought (according to PDSI) was found to be a significant predictor of cholera outbreaks, with drier conditions seeing higher cholera outbreak occurrence. While previous studies have implicated drought in cholera outbreaks [2,10,11], our study models drought in isolation allowing a more in-depth investigation of its impacts, which have been largely understudied in comparison to flooding. In addition, we tested whether drought is likely to influence cholera outbreaks under scenarios of climatic change and socio-economic development (attainment of the SDGs). While we found drought will continue to be an important hazard for cholera outbreaks in the future, our results suggest that gains in sustainable development (reduction of poverty, increased water security) may offset cholera risk in the future.

Temperature was identified as a significant predictor, providing another link between changing drought risk and increased cholera outbreak occurrence, as an increased temperature is important in both drought onset and duration. The positive relationship between temperature and cholera is expected, as cholera is considered a temperature-sensitive pathogen, with optimum growth at elevated temperatures (up to a threshold) [46]. This may also represent an independent effect of temperature from drought and why both variables are independently selected in the model. For example, a 1°C rise in temperature was associated with a 2-fold increase in cholera cases in Zanzibar [8]. Moreover, when run in the univariate models, precipitation had a slightly negative coefficient, again providing a potential link between drought, decreased water availability and cholera outbreaks. Precipitation, however, was not selected in the final model, potentially suggesting that precipitation effects for cholera in Africa, may be less important than temperature.

The inclusion of more than one type of drought index in the best-fit model (PDSI and water withdrawal) shows the importance of considering several drought definitions and measures when investigating its implications. Drought is a complex phenomenon involving climate, agriculture, water stress and societal response and therefore including additional drought variables can help capture the varying elements of the hazard, exposure and vulnerability. Water withdrawal per capita was a highly significant environmental variable in the model, linking to the original hypothesis that a reduction in water availability leads to riskier water practices. More water withdrawal suggests higher water availability for drinking and washing and a reduction in risky behaviour such as with multi-use water. Better water management may help mitigate negative drought-related health outcomes, and when water is available, this should not be exploited to avoid times of scarcity.

Cholera is a disease of inequity and poverty and is often seen in combination with poor WASH facilities [14,49]. Here, poverty was the most significant variable (according to the p values) included in the model and may suggest that environmental determinants of cholera are only key drivers up to certain thresholds and then socio-economic covariates are more appropriate predictors [13]. For example, droughts have been known to impact the US and Europe [47,48], but large-scale cholera outbreaks do not occur due to generally high levels of sanitation and hygiene. Several socio-economic covariates were expected to be important here but only poverty was selected in the final model and all socioeconomic covariates were independently selected for model inclusion. A possible explanation is that other socio-economic covariates such as, sanitation, hygiene, drinking water and people living in informal settlements is captured within the effects of poverty and possibly enhancing its impact. Even with the ideal environment for cholera to proliferate, social conditions allow the link to be made for pathogen exposure and spread. Poor access to WASH facilities means that large groups of people are at risk, not just for cholera, but for several other diseases. For example, nearly 90% of diarrhoeal disease has been attributed to sub-optimal WASH [50]. These findings highlight the need to meet or exceed the SDGs, lifting people out of poverty and providing basic sanitation and hygiene as a public health priority.

The scenario dataset and projections provide some insight into the future importance of climate and socio-economic development on cholera outbreak occurrence in Africa. Historical and projected changes are spatially heterogenous but projected continental trends under S3 slightly increased cholera outbreak occurrence to 2070. Whereas, under S2 and S1 cholera occurrence decreased to 2070, with S1 showing the lowest levels. The projected changes over the next 50 years show that reducing poverty, expanding sustainable freshwater availability and striving for greater emissions reductions will be important for achieving positive health outcomes. How societies will continue to respond and adapt to climate change and drought is difficult to determine in the future and therefore understanding future risks can be challenging. As with any projections and the creation of scenarios, uncertainty can be high, arising from theoretical, methodological and computational challenges in projecting future climate change and its consequences. There are also the realities of meeting or exceeding the SDGs. Several of the terms within the SDGs are ambiguous, consequently the aims and roadmap to achieve them are not clearly defined. Finally, we did not consider that in a “worst-case” scenario poverty and water withdrawal may regress or the introduction of new strains and changing immunity could complicate cholera eradication efforts. Despite this, with decreasing poverty and the expansion of freshwater availability, even the introduction of new cholera cases and strains could be offset.

Climate, drought and socio-economic data were missing for several countries and years, meaning that data had to be averaged or omitted. This meant that data were then missing from the model, or assumptions had to be made both spatially and temporally about conditions in certain countries, potentially introducing error. Cholera is largely underreported, and many people never seek formal medical assistance. The WHO’s most optimistic estimate suggests only 5-10% of cases are reported [51], possibly due to a spectrum of transmission dynamics and lulls in cases meaning focus on tracking the diseases can be lost [52]. Countries can be disincentivized to report outbreaks due to potential impact on tourism and trade [53]. Considering this underreporting, issues may have arisen from assigning the outcome variable to zero for missing years, as this could have led to the underrepresentation of cholera outbreaks. Given the results of the sensitivity analysis in Supplementary Information 1, however, we believe this is the best interpretation of missing values, as removing values created issues when trying to select covariates from small numbers of data points. Furthermore, the cholera data lack age and sex-disaggregation, meaning that demographic differences were not captured. GLMs assume a monotonic relationship and therefore non-linear effects of several covariates might not be captured and evaluating these non-linear effects are a potential area of future work. This issue may also have been present for the S3 drought projections as some countries fit the linear trend better than others.

## Conclusions

In conclusion, the relationships between temperature, drought and water withdrawal help add further evidence to the original hypothesis that hotter and drier conditions and a lack of freshwater availability increases cholera outbreak occurrence, potentially through risky water behaviours. Although elevated pathogen concentrations are difficult to distinguish from these results, the importance of elevated temperatures and its effect on cholera may be related to increases in pathogen concentrations. Socio-economic variables came out highly significant in the best-fit model, showing the impact of vulnerability in times of water shortage and the importance of lifting people out of poverty to improve health and reduce mortality.

The work presented here offers additional insight into how climate change may yield health impacts in the future and work should build on these results, to understand these relationships on a finer spatial scale. High burden countries such as the DRC and Nigeria saw very few changes in cholera over the projected period and scenarios, showing potential areas for further work to understand outbreak drivers and mitigators in the most at-risk countries.

## Data Availability

All the data used in this analysis are publicly available and cited throughout the manuscript.

## List of abbreviations

WHO: World Health Organization
WASH: Water, sanitation and hygiene
PDSI: Palmers Drought Severity Index
PET: Potential evapotranspiration
GLM: Generliased linear model
BIC: Bayesian Information Criterion
AUC: Area under the receiver operator characteristic curve
AIC: Akaike Information Criterion
SDG: Sustainable Development Goals
LOO: Leave-one-out cross validation
DRC: Democratic Republic of Congo

## Declarations

### Ethical approval and consent to participate

This manuscript does include human data, but no ethical approval or consent was needed, as this data is freely available through WHO, the WorldBank and the UN Development Programme and completely anonymised. No animal data was used in this research.

### Consent for publication

Not applicable

### Availability of data and materials

The datasets used in this research are all publicly available and referenced throughout the manuscript.

### Authors’ contributions

GECC was part of the study design and conceptualisation of ideas, collected the data and ran the analysis, wrote and finalised the manuscript and incorporated any feedback. IK provided expertise on droughts, health and climate change and revised several drafts. NG provided support in the statistical analysis. WH extracted the environmental data from WorldClim and provided knowledge on the data. KAMG was part of the study design and conceptualisation of ideas, provided expertise in the statistical analysis and revised several drafts. KAM was part of the study design and conceptualisation of ideas, provided expertise on cholera, ecology and climate change, provided supervision and revised several drafts.

## Acknowledgments

We would like to thank those who collected and curated the publicly available datasets used here.

